# Unraveling COVID-19 Relationship with Anxiety Disorders and Symptoms

**DOI:** 10.1101/2023.07.21.23293001

**Authors:** Zeynep Asgel, Manuela R. Kouakou, Dora Koller, Gita A. Pathak, Brenda Cabrera-Mendoza, Renato Polimanti

**Affiliations:** Department of Psychiatry, Yale University School of Medicine, New Haven, CT, USA; Department of Genetics, Microbiology, and Statistics, Faculty of Biology, University of Barcelona, Catalonia, Spain; VA Connecticut Healthcare System, West Haven, CT, USA; Wu Tsai Institute, Yale University, New Haven, CT, USA

## Abstract

**Background:** While COVID-19 outcomes are associated with increased anxiety, individuals affected by anxiety disorders are more likely to develop severe COVID-19 outcomes.

**Methods:** We used genome-wide data from UK Biobank (up to 420,531 participants), FinnGen Project (up to 329,077 participants), Million Veteran Program (175,163 participants), and COVID-19 Host Genetics Initiative (up to 122,616 cases and 2,475,240 controls) to investigate possible causal effects and shared genetic mechanisms linking COVID-19 outcomes to anxiety disorders and symptoms.

**Results:** We observed a strong genetic correlation of anxiety disorder with COVID-19 positive status (rg=0.35, p=2 × 10^-4^) and COVID-19 hospitalization (rg=0.31, p=7.2 × 10^-4^). Among anxiety symptoms, “Tense, sore, or aching muscles during worst period of anxiety” was genetically correlated with COVID-19 positive status (rg=0.33, p=0.001), while “Frequent trouble falling or staying asleep during worst period of anxiety” was genetically correlated with COVID-19 hospitalization (rg=0.24, p=0.004). Through a latent causal variable analysis, we observed that COVID-19 outcomes have statistically significant genetic causality proportion (gcp) on anxiety symptoms (e.g., COVID-19 positive status→“Recent easy annoyance or irritability” │gcp│=0.18, p=6.72 × 10^-17^). Conversely, anxiety disorders appear to have a possible causal effect on COVID-19 (│gcp│=0.38, p=3.17 × 10^-9^). Additionally, we also identified multiple loci with evidence of local genetic correlation between anxiety and COVID-19. These appear to be related to genetic effects shared with lung function, brain morphology, alcohol and tobacco use, and hematologic parameters.

**Conclusions:** This study provided important insights into the relationship between COVID-19 and mental health, differentiating the dynamics linking anxiety disorders to COVID-19 from the effect of COVID-19 on anxiety symptoms.

## INTRODUCTION

The COVID-19 pandemic has had a profound impact on mental health. Several challenges associated with the pandemic, such as social isolation, economic uncertainty and fear of illness led to an increase in negative mental health outcomes (Fitzpatrick, Harris, & Drawve, 2020; Kampfen et al., 2020), especially those related to anxiety disorders and symptoms (Choi, Hui, & Wan, 2020; Pierce et al., 2020; Verma & Mishra, 2020). This increase can mainly be observed in individuals that have been hospitalized due to COVID-19 and in individuals with a history of mental illness (Liu, Erdei, & Mittal, 2021; Mazza et al., 2020). Clinical factors such as length of hospital stay, need for mechanical ventilation, and comorbidities are likely to contribute to COVID-19 and anxiety association (Mazza et al., 2020). Anxiety also leads to an increased risk of neurological and psychiatric disorders in individuals who have survived COVID-19 (Taquet, Geddes, Husain, Luciano, & Harrison, 2021). Beyond brain-related conditions, the relationship between anxiety and COVID-19 can also impact the immune system and increase the risk of infections by altering innate and adaptive immunity, resulting in a compromised host defense system, and increased systemic inflammation (Mazza et al., 2020).

Advances in genetics research are providing new tools to investigate the genetic architecture of psychopathology and its relationship with other health outcomes (Andreassen, Hindley, Frei, & Smeland, 2023; Demkow & Wolanczyk, 2017). By identifying genetic variants associated with both psychiatric disorders and somatic conditions, we can gain a better understanding of the epidemiological relationships and biological mechanisms involved in these comorbidities.

Genetic predisposition significantly contributes to the development of psychiatric disorders (Sullivan & Geschwind, 2019). In particular, twin-based studies showed that genetic factors account for 20 to 60% of the phenotypic variance across anxiety subtypes (Ask et al., 2021). Loci associated with anxiety disorders and traits also affect other health outcomes related to somatic comorbidities, such as gastrointestinal alterations (Eijsbouts et al., 2021), the immune system (Werner et al., 2022), blood pressure (Thorp et al., 2021), pain (Pinheiro, Morosoli, Colodro-Conde, Ferreira, & Ordonana, 2018), and insomnia symptoms (Lane et al., 2017).

Genetically informed studies also highlighted potential causal relationships linking COVID-19 to different psychiatric disorders (W. Chen et al., 2022; N. Liu et al., 2021; Luykx & Lin, 2021).

While the results of these studies provided insights into possible cause-effect relationships, observational studies highlighted a complex interplay between COVID-19 and anxiety where bidirectional effects and shared pathogenesis could both play a role (Young et al., 2022; Zhang et al., 2023). Thus, a more comprehensive analysis of COVID-19 comorbidity with anxiety disorders and traits can shed light on genetic mechanisms that may be important in developing targeted interventions and improving mental health outcomes. Leveraging large-scale datasets from UK Biobank (UKB) (Bycroft et al., 2018), FinnGen Project (Kurki et al., 2023), Million Veteran Program (MVP) (Gaziano et al., 2016), and COVID-19 Host Genetics Initiative (HGI) (Initiative, 2021), we investigate the COVID-19 relationship with anxiety disorders and traits, testing multiple pleiotropic mechanisms that may be responsible for the comorbidity observed.

## METHODS

### Data Sources

We investigated data generated from large-scale genome-wide association studies (GWAS) of anxiety disorders and traits and COVID-19 outcomes.

With respect to anxiety-related phenotypes, we analyzed genome-wide association statistics from previous UKB (Pan-UKB team, 2020), FinnGen Project (Kurki et al., 2023), and MVP analyses (Gaziano et al., 2016). For UKB, GWAS data were generated by the Pan-UKB analysis as previously described (Pan-UKB team, 2020). Briefly, the genome-wide association analysis was conducted using regression models available in Hail (available at https://github.com/hail-is/hail) and including the top-20 within-ancestry principal components (PC), sex, age, age^2^, sex×age, and sex×age^2^ as covariates. Based on SNP-based heritability (SNP-h2) z score > 4 (see *SNP-based Heritability and Genetic Correlation* section), we selected anxiety phenotypes (up to 420,531 participants of European descent) derived from UKB electronic health records and questionnaires (Supplementary Table 1). As described previously, the FinnGen GWAS Release 8 data (FinnGen Project, 2022) were generated applying REGENIE regression model (Mbatchou et al., 2021) and including age, sex, top-10 within-ancestry 10 PCs, and genotyping batch as covariates. For FinnGen, anxiety endpoints (up to 329,077 participants of European descent) were derived from electronic health records (Supplementary Table 2). With respect to MVP, we investigated data generated from a previous GWAS of anxiety symptoms assessed via the Generalized Anxiety Disorder (GAD) 2-item scale in 175,163 participants of European descent (Levey et al., 2020).

Genome-wide association statistics were also obtained from the COVID-19 Host Genetics Initiative (HGI) (Initiative, 2021). In the present study, we analyzed Release 7 GWAS data (The COVID-19 Host Genetics Initiative, 2023) related to three outcomes: “very severe respiratory confirmed covid vs. population” (13,769 cases and 1,072,442 controls of European descent); “hospitalized covid vs. population” (32,519 cases and 2,062,805 controls of European descent); “covid vs. population” (122,616 cases and 2,475,240 controls of European descent). COVID-19 HGI GWAS were based on large meta-analyses of multiple cohorts, including UKB, FinnGen, and MVP. For this reason, we investigated the pleiotropy between COVID-19 and anxiety using methods that can account for sample overlap among the GWAS datasets investigated.

Unfortunately, we had to limit our analyses on GWAS conducted in individuals of European descent, because no large-scale anxiety and COVID-19 GWAS were available for other ancestry groups.

### SNP-based Heritability and Genetic Correlation

To estimate SNP-h2 and genetic correlation among COVID-19 outcomes and anxiety-related traits, the linkage disequilibrium score regression (LDSC) method (Bulik-Sullivan et al., 2015; Gazal et al., 2017) was applied. The LD scores were pre-computed based on 1000 Genomes Project Phase 3reference data on individuals of European ancestry. Following the LDSC developer’s recommendations (Bulik-Sullivan et al., 2015), the genetic correlation analysis was limited to phenotypes with SNP-h2 z score>4. To account for the number of tests performed in the genetic correlation analysis, we applied a false discovery rate correction (FDR q<0.05).

### Genetically Informed Causal Inference Analysis

The latent causal variable (LCV) method (O’Connor & Price, 2018) was used to test whether the genetic correlation observed is due to possible causal effects. Assuming a single effect-size distribution, the LCV method examines a latent trait between genetically correlated traits to estimate the proportion of genetic causality (gĉp) that ranges from 0 to 1. Partial causality is indicated by values close to zero, while full causality is indicated by values close to 1. The direction of the putative causal effect is reflected by positive and negative gĉp values, and the LCV genetic correlation estimate (rho) provides information on whether the LCV effect is positive or negative. The analysis was performed using the European-ancestry LD scores in R.

### Local Genetic Correlation Analysis

To identify the genomic regions contributing to pathogenic mechanisms shared between COVID-19 and anxiety-related phenotypes, the local analysis of [co]variant association (LAVA) (Werme, van der Sluis, Posthuma, & de Leeuw, 2022) method was used. To account for correlated SNPs due to LD, LAVA converts the marginal SNP effects into joint effects using an external LD reference panel, which in our study was derived from the 1000 Genomes Project European populations. The genome was divided into roughly 2,495 semi-independent blocks of approximately 1Mb to create regions. LAVA also uses the intercept from bivariate LDSC to address any potential sample overlap. To account for the number of tests performed in the local genetic correlation analysis, we applied FDR correction (FDR q<0.05).

### Colocalization Analysis

Hypothesis Prioritization in multi-trait Colocalization (HyPrColoc) (Foley et al., 2021) analysis was performed with respect to the regions that showed FDR-significant local genetic correlation to identify specific loci that may be related to both COVID-19 outcomes and anxiety-related phenotypes via tissue-specific transcriptomic regulation. Genotype-Tissue Expression (GTEx) v8 (GTEx Consortium, 2020) was used to derive information regarding expression quantitative trait loci (eQTL) in the regions identified in the LAVA analysis. Following HyPrColoc developers’ recommendation (Foley et al., 2021), we defined COVID-19-anxiety colocalized loci as those with a posterior probability >70%.

## RESULTS

### SNP-based Heritability and Genetic Correlation

COVID-19 outcomes and multiple anxiety phenotypes showed a statistically significant SNP-h2 (z-score>4; Table 1) with evidence of a consistent genetic correlation between these two health domains (Figure 1; Supplemental Table 3). Specifically, 20 pairwise combinations survived FDR multiple testing correction (FDR q<0.05). While only “Seen general practitioner for nerves, anxiety, tension or depression” was genetically correlated with “very severe respiratory confirmed covid” outcome (rg=0.14, p=9 × 10^-4^), “hospitalized covid” and “covid” were genetically correlated with multiple anxiety phenotypes (Supplemental Table 3). For both, anxiety disorder (phecode 300.1) showed a high genetic correlation with “hospitalized covid” (rg=0.31, p=7.2 × 10^-4^) and “covid” (rg=0.35, p=2 × 10^-4^).

**Figure 1.**
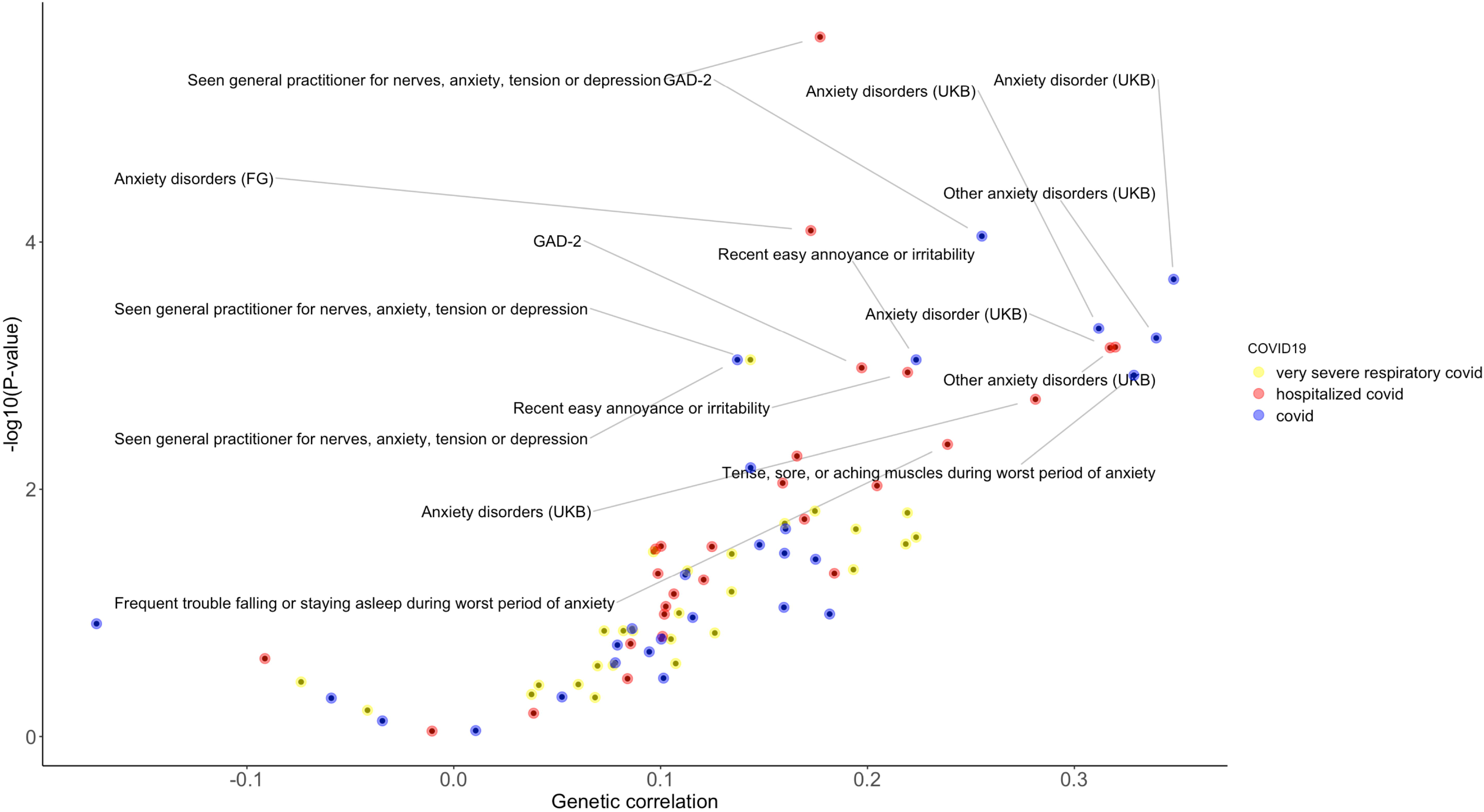
Genetic correlations of COVID-19 outcomes with anxiety phenotypes assessed in UK Biobank (UKB), FinnGen Study (FG), and Million Veteran Program (MVP). Labels are reported for genetic correlation surviving false discovery rate multiple testing correction. Full results are available in Supplemental Table 3.

**Table 1.** COVID-19 and anxiety phenotypes with SNP-h2 z>4. Sample size (N) is reported as case-control for binary traits and as number of individuals for quantitative traits. Additional information regarding UK Biobank (UKB) and FinnGen phenotypes are available in Supplemental Tables 1 and 2, respectively.

| Phenotype | Source | N | SNP-h2 Z |
| --- | --- | --- | --- |
| very severe respiratory confirmed covid vs. population | COVID-19<br>HGI | 13,769-1,072,442 | 5.39 |
| hospitalized covid vs. population |  | 32,519-2,062,805 | 6.67 |
| covid vs. population |  | 122,616-2,475,240 | 6.67 |
| GAD-2 total score | MVP | 175,163 | 13.61 |
| Anxiety disorders (phecode: 300) | UKB | 10,449-369,930 | 8.13 |
| Anxiety disorder (phecode: 300.1) |  | 9,705-369,930 | 7.57 |
| Other anxiety disorders (ICD-10: F41) |  | 9,006-411,525 | 6.65 |
| Non-cancer illness code, self-reported: anxiety/panic attacks |  | 5,800-414,673 | 5.51 |
| Tense, sore, or aching muscles during worst period of anxiety |  | 15,013-23,267 | 5.24 |
| Frequent trouble falling or staying asleep during worst period of anxiety |  | 33,529-7,403 | 4.84 |
| Mental health problems ever diagnosed by a professional: Anxiety, nerves or generalized anxiety disorder |  | 19,338-116,659 | 8.84 |
| Substances taken for anxiety: Medication prescribed to you (for at least two weeks) |  | 14,874-99,435 | 10.36 |
| Activities undertaken to treat anxiety: Talking therapies, such as psychotherapy, counselling, group therapy or CBT |  | 14,714-99,595 | 8.70 |
| Activities undertaken to treat anxiety: Other therapeutic activities such as mindfulness, yoga or art classes |  | 6,557-107,752 | 6.46 |
| Seen general practitioner for nerves, anxiety, tension or depression |  | 144,320 273,287 | 22.71 |
| Seen a psychiatrist for nerves, anxiety, tension or depression |  | 47,877-370,757 | 15.30 |
| Impact on normal roles during worst period of anxiety |  | 41,979 | 4.58 |
| Recent easy annoyance or irritability |  | 135,686 | 10.91 |
| Recent feelings or nervousness or anxiety |  | 135,825 | 9.82 |
| Recent inability to stop or control worrying |  | 135,808 | 8.87 |
| Recent feelings of foreboding |  | 135,687 | 10.90 |
| Recent trouble relaxing |  | 135,937 | 10.83 |
| Recent restlessness |  | 136,022 | 8.62 |
| Recent worrying too much about different things |  | 135,794 | 10.31 |
| Frequency of inability to stop worrying during worst period of anxiety |  | 41,680 | 4.70 |
| Generalized anxiety disorder (F5_GAD) | FinnGen | 3,923-307,558 | 4.77 |
| All anxiety disorders (F5_ALLANXIOUS) |  | 21,519-307,558 | 12.89 |
| Phobic anxiety disorders (F5_PHOBANX) |  | 3,791-307,558 | 6.23 |
| Other anxiety disorders (F5_ANXIETY) |  | 14,707-307,558 | 12.4 |
| Anxiety disorders (ANXIETY_EXMORE) |  | 35,385-254,976 | 15.09 |

### Genetically Informed Causal Inference Analysis

The LCV analysis showed that some of the genetic correlations observed between COVID-19 outcomes and anxiety phenotypes were related to possible causal relationships (Supplemental Table 4). After applying FDR multiple testing correction (FDR q<0.05), statistically significant gcp estimates for nine of the pairwise combinations tested (Figure 2). As mentioned in the method section, positive and negative gcp values reflect the direction of the putative causal effect (i.e., COVID-19 → anxiety and anxiety → COVID-19, respectively) while the sign of the effect is given by the rho statistics. With respect to FDR-significant negative gcp results, there were four anxiety phenotypes potentially affecting “covid” outcome. Three of them were related to case-control definitions derived from UKB electronic health records and the most statistically significant result was related to anxiety disorder (phecode 300.1 gcp=-0.38, p=3.17 × 10^-9^; rho=0.35, SE=0.1). The other anxiety phenotype affecting “covid” outcome was “Tense, sore, or aching muscles during worst period of anxiety” (gcp=-0.21, p=6.58 × 10^-4^; rho=0.26, SE=0.1). With respect to FDR-significant positive gcp results, “covid” and “very severe respiratory confirmed covid” outcomes showed consistent effects across several anxiety symptoms with the strongest statistical evidence being “Recent easy annoyance or irritability” (gcp=0.18, p=6.72 × 10^-17^; rho=0.23, SE=0.07; gcp=0.1, p=1.42 × 10^-9^; rho=0.15, SE=0.07, respectively).

**Figure 2.**
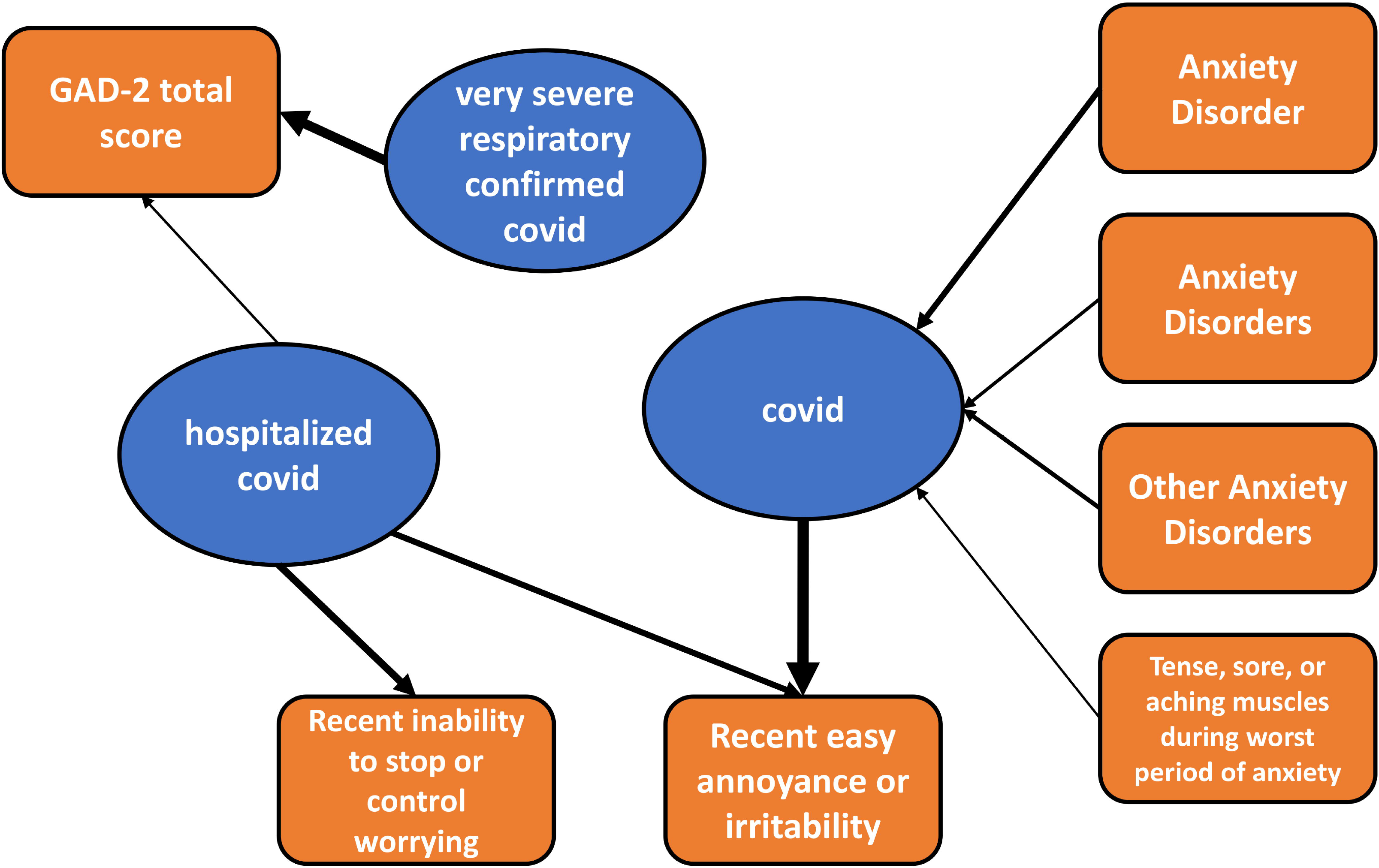
Latent causal variable network showing the effects between COVID-19 and anxiety phenotypes. The arrow width corresponds to the statistical significance of the genetic causal proportion. Full results are available in Supplemental Table 4.

### Local Genetic Correlation Analysis

We identified eight local genetic correlations between COVID-19 outcomes and anxiety phenotypes after FDR-multiple testing correction (FDR q<0.05; Table 2; Supplemental Table 5). With respect to case-control definitions of anxiety disorders based on UKB electronic health records, we observed multiple results for two loci. One of them was specifically related to “hospitalized covid” (7:130,418,705-131,856,481; e.g., anxiety disorder, phecode 300.1: rho=1, p=8.02 × 10^-7^; Figure 3), while the other locus showed multiple local genetic correlations with “covid” (2:234,945,578-235,625,789; e.g., anxiety disorder, phecode 300.1: rho=1, p=5.48 × 10^-6^). Similar to the other analyses, most of the local genetic correlations were related to “hospitalized covid” and “covid” outcomes. However, we also observed a local genetic correlation between “very severe respiratory confirmed covid” and GAD-2 score (15:33,484,268-34,410,072; rho=0.92, p=7.35 × 10^-6^). Additionally, in line with the overall positive genetic correlation between COVID-19 outcomes and anxiety phenotypes, local genetic correlations were also positive with the only exception being the “22:27,192,924-27,952,441” locus for “covid” and “recent inability to stop or control worrying” (rho=1, p=5.48 × 10^-6^).

**Figure 3.**
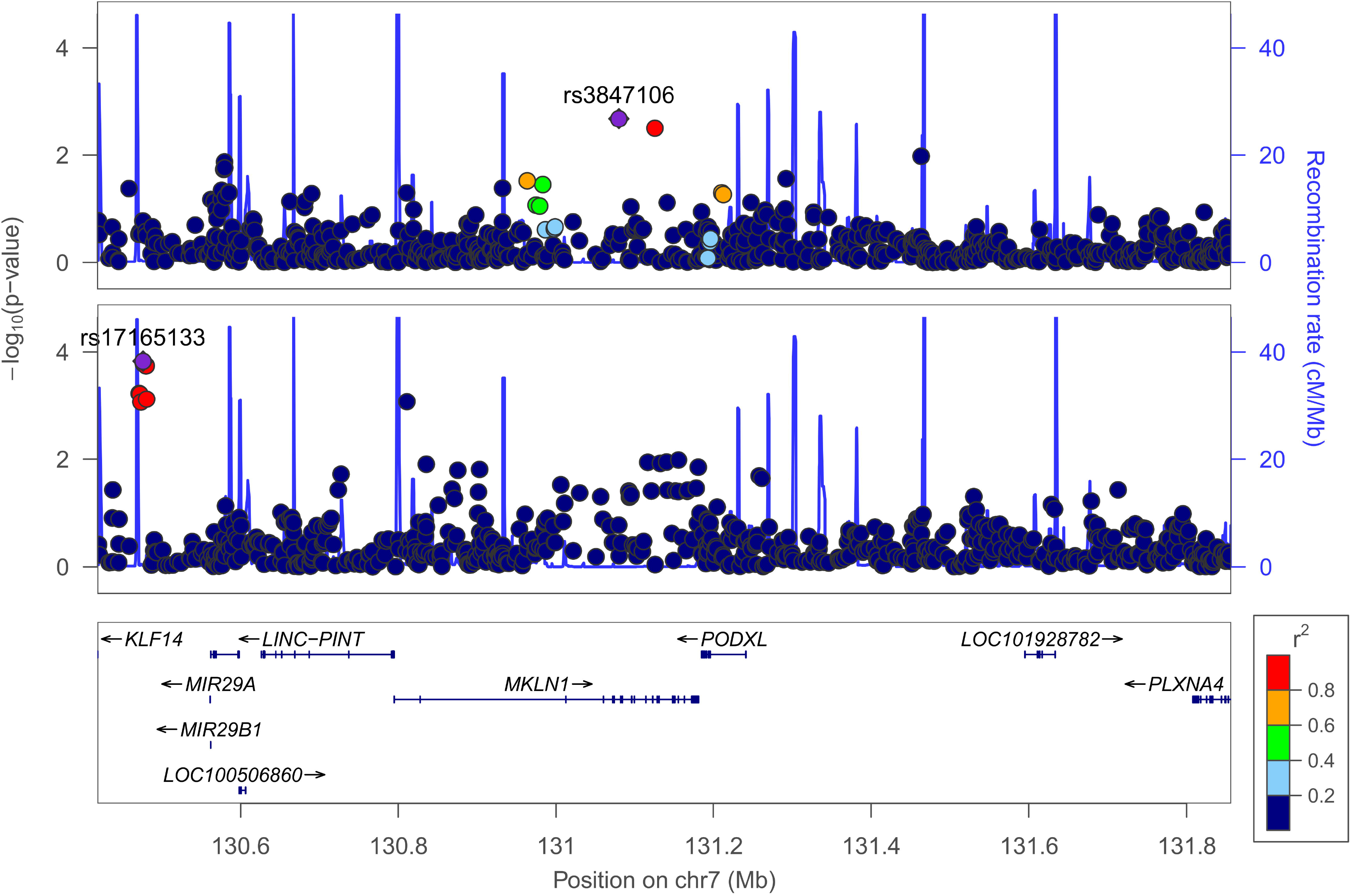
Regional Manhattan plot of “7:130,418,705-131,856,481” locus with respect to anxiety disorder (phecode 300.1; bottom panel) and “hospitalized covid” (upper panel).

**Table 2.** Local genetic correlations between COVID-19 outcomes and anxiety phenotypes surviving FDR multiple testing correction (FDR q<0.05). Full local genetic correlation results are available in Supplemental Table 5. Details regarding genome-wide significant associations reported in the GWAS catalog for each locus are available in Supplemental Table 6, 7, 8, 9, and 10.

| Locus<br>(chromosome:position) | COVID-19 Phenotype | Anxiety Phenotype | rho | p | GWAS Catalog<br>Associations, N |
| --- | --- | --- | --- | --- | --- |
| 15:33,484,268-34,410,072 | very severe respiratory<br>confirmed covid | GAD-2 score (MVP) | 0.92 | 7.35E-06 | 22 |
| 7:130,418,705-131,856,481 | hospitalized covid | Anxiety disorders (phecode 300) | 1 | 1.08E-07 | 508 |
|  |  | Anxiety disorder (phecode 300.1) | 1 | 8.02E-07 |  |
|  |  | Anxiety disorders (ICD10: F41) | 1 | 3.66E-06 |  |
| 9:14,244,748-14,902,924 |  | Tense, sore, or aching muscles<br>during worst period of anxiety | 0.88 | 4.18E-06 | 86 |
| 2:234,945,578-235,625,789 | covid | Anxiety disorders (phecode 300) | 1 | 1.49E-06 | 34 |
|  |  | Anxiety disorder (phecode 300.1) | 1 | 5.48E-06 |  |
| 22:27,192,924-27,952,441 |  | Recent inability to stop or control<br>worrying | -0.92 | 1.02E-05 | 95 |

Within the loci with FDR-significant local genetic correlation, the multi-trait analysis did not identify a single variant with evidence of colocalization among genetic effects related to COVID-19 outcomes, anxiety phenotypes, and tissue-specific transcriptomic regulation (posterior probability <70%).

To characterize the possible phenotypic implications related to the loci identified by the local genetic correlation analysis, we investigated the genome-wide significant associations (GWS, p<5 × 10^-8^) available from the GWAS catalog (Sollis et al., 2023). A total of 508 GWS associations were reported within “7:130,418,705-131,856,481” locus (Supplemental Table 6) with the strongest ones being insulin-like growth factor 1 (IGF-1) levels (rs157934 p=2 × 10^-109^) and high-density lipoprotein cholesterol levels (rs13234269 p=5 × 10^-102^). Many GWS associations were also identified within “22:27,192,924-27,952,441” locus (N=95; Supplemental Table 7) with lung function (rs134041 p=1 × 10^-27^) and brain-imaging phenotypes (rs134016 p=8 × 10^-19^) among the most significant traits. GWS associations with traits potentially relevant for anxiety-COVID19 comorbidity were also observed within the “9:14,244,748-14,902,924” locus (Supplemental Table 8; e.g., educational attainment rs10124571 p=8 × 10^-19^, smoking initiation rs10810153 p=3 × 10^-18^, and brain morphology rs1323342 p=4 × 10^-12^). The “2:234,945,578-235,625,789” locus was characterized by GWS associations with multiple hematological parameters (Supplemental Table 9; e.g., mean corpuscular volume rs1356334 p=2 × 10^-36^, red cell distribution width rs1517006 p=1 × 10^-26^, and platelet count rs114266592 p=2 × 10^-15^). GWS associations with educational attainment (rs35483472 p=3 × 10^-12^), alcohol consumption (rs117799466 p=3 × 10^-9^), mean platelet volume (rs8039126 p=2 × 10^-11^), and hepatic protein levels (rs78319531 p=3 × 10^-8^) were observed within the “15:33,484,268- 34,410,072” locus (Supplemental Table 10).

## DISCUSSION

The COVID-19 pandemic had a profound impact on mental health, leading to a substantial increase in anxiety-related conditions (Choi et al., 2020; Pierce et al., 2020; Verma & Mishra, 2020). However, the dynamics underlying this relationship are still unclear. Leveraging large-scale genome-wide datasets, the current study aimed to understand the possible causal effects and shared genetic mechanisms linking COVID-19 outcomes with anxiety disorders and traits.

The genetic correlation analysis showed a consistent overlap of COVID-19 outcomes with anxiety disorders and symptoms derived from different definitions based on electronic health records or self-reported information. This supports COVID-19 association across the anxiety spectrum rather than being specific to a certain anxiety domain. Indeed, we observed genetic correlations of COVID-19 outcomes with both behavioral symptoms (e.g., “Recent inability to stop or control worrying”) and physical symptoms (e.g., “Tense, sore, or aching muscles during worst period of anxiety”). However, the LCV analysis showed that the genetic overlap of COVID-19 across anxiety spectrum may be due to two distinct paths. Anxiety disorders and anxiety physical symptoms may have a causal effect on COVID-19, while COVID-19 may play a causal role on anxiety behavioral symptoms. Additionally, worse COVID-19 outcomes (i.e., “very severe respiratory confirmed covid” and “hospitalized covid”) appear to affect overall anxiety severity assessed via the GAD-2 scale, which is based on behavioral symptoms (Sapra, Bhandari, Sharma, Chanpura, & Lopp, 2020). The possible role of anxiety disorders on COVID- 19 susceptibility is in line with a cross-sectional study of 2,535,098 participants that showed increased odds of COVID-19-related mortality among patients affected by anxiety disorders (Teixeira et al., 2021). With respect to COVID-19 consequences, a longitudinal study of 54,442 participants reported an association of COVID-19 with subsequent anxiety symptoms that did not interact with pre-pandemic mental health (Thompson et al., 2022).

Beyond possible cause-effect relationships, our local genetic correlation analysis identified specific loci that could play important roles in the pleiotropic mechanisms linking COVID-19 outcomes to anxiety disorders and symptoms. The subsequent colocalization analysis showed that the local genetic correlations observed were not due to the effect of a single variant. This suggests that the effect of multiple variants within each locus may be responsible for the local genetic correlations observed. In line with this hypothesis, we observed that multiple GWS associations were reported by previous studies in the loci identified. In “7:130,418,705- 131,856,481” locus, the strongest GWS association was related to IGF-1 levels (Sinnott-Armstrong et al., 2021). A Mendelian randomization analysis showed that genetically predicted high IGF-1 levels are protective with respect to COVID-19 susceptibility and hospitalization (Li et al., 2022). There is also evidence from human and animal experiments that circulating IGF-1 modulates mood and can reflect stress vulnerability (Santi, Bot, Aleman, Penninx, & Aleman, 2018). In the same locus, there are multiple GWS associations with lipid levels (Graham et al., 2021). Multiple studies highlighted the relationship of COVID-19 with lipid profile and the substantial implications for disease severity and prognosis (Surma, Banach, & Lewek, 2021).

Similarly, altered lipid metabolism has been hypothesized to affect anxiety disorders through multiple pathways, such as oxidative stress and inflammation (Humer, Pieh, & Probst, 2020). Multiple GWS associations were also present in “22:27,192,924-27,952,441” locus. Several of them were related to lung function (Kichaev et al., 2019) and pulmonary diseases, such as chronic obstructive pulmonary disease (Cosentino et al., 2023). Other GWS associations were also to multiple hematological parameters (e.g., mean corpuscular volume, mean corpuscular hemoglobin, and platelet count) (M. H. Chen et al., 2020). In this locus, there was also a GWS association with the grey matter volume in vermis crus II of the cerebellum (Smith et al., 2021). This region has been associated with social mentalizing and emotional self-experiences (Van Overwalle, Ma, & Heleven, 2020). The relationship of hematological parameters with the local genetic correlation between COVID-19 and anxiety is also supported by another of the loci identified, “2:234,945,578-235,625,789”, where many GWAS associations with blood-related traits have been reported (e.g., mean corpuscular volume, red cell distribution width, mean corpuscular hemoglobin) (M. H. Chen et al., 2020). Altered hematological profile has been previously linked to negative COVID-19 outcomes (Mei et al., 2020) and anxiety symptoms (Shafiee et al., 2017). Convergent findings were also observed in “9:14,244,748-14,902,924” and “15:33,484,268-34,410,072” loci, where multiple GWS associations were reported for educational attainment (Okbay et al., 2022), alcohol consumption (Cole, Florez, & Hirschhorn, 2020), and smoking initiation (Saunders et al., 2022). These factors have been associated with COVID-19 susceptibility in observational studies of diverse cohorts (Matthay et al., 2022; Schafer, Santos, Quadra, Dumith, & Meller, 2022). The relationship of anxiety symptoms and disorders with academic achievements has been reported by multiple investigations (Brumariu, Waslin, Gastelle, Kochendorfer, & Kerns, 2022). Similarly, there is a well-known association of alcohol consumption and tobacco smoking with anxiety disorders and symptoms (Anker & Kushner, 2019; Fluharty, Taylor, Grabski, & Munafo, 2017). Taken together, the GWS associations reported within loci identified by the LAVA analysis suggest that genetic effects related to other health outcomes may be responsible for the local genetic correlation observed between COVID-19 and anxiety.

The current study has some limitations. The difference in the statistical power of the GWAS investigated may be responsible for some of the null results observed. In particular, most of our results were related to “hospitalized covid” and “covid” phenotypes. The limited sample of the “very severe respiratory confirmed covid” GWAS likely affected the statistical power to investigate the relationship between this COVID-19 outcome and anxiety. Another important limitation is related to the fact that our study analyzed only genome-wide data generated from individuals of European descent, because of the lack of COVID-19 and anxiety GWAS representative of other worldwide populations. Accordingly, our findings may not be generalizable to other ancestry groups. Similarly, there may be also important differences between sexes that are not observable in the analyses performed.

In conclusion, this study provided important insights into the relationship between COVID-19 and anxiety, highlighting the possible contribution of multiple pleiotropic paths. Specifically, Anxiety disorders may increase COVID-19 susceptibility, while COVID-19 outcomes may contribute to anxiety behavioral symptoms. Additionally, genetic effects shared with risk factors may also play an important role in the association between COVID-19 and anxiety.

## Data Availability

All data produced in the present work are contained in the manuscript.

## Acknowledgments

The authors thank the participants and the investigators involved in the UK Biobank, FinnGen Project, Million Veteran Program, and COVID-19 Host Genetics Initiative.

## Financial support

This study was supported by grants from the National Institutes of Health (RF1 MH132337 and R33 DA047527) and One Mind. The funding sources had no role in the design of this study, its execution, analyses, interpretation of the data, and the decision to publish the results.

## Financial Disclosures

RP received a research grant from Alkermes and is paid for his editorial work on the journal Complex Psychiatry.

## Notes

### Author Declarations

The study was conducted using publicly available genome-wide association statistics. Details regarding the data location are provided in the manuscript.

